# AutoMRAI: A Multi-Omics Causal Inference Platform Using Structural Equation Modelling

**DOI:** 10.1101/2025.11.17.25340455

**Authors:** Fadhaa Ali

## Abstract

The integration of causal inference, artificial intelligence (AI), and multi-omics data represents a transformative frontier for unravelling the complex mechanisms underlying health and disease. Traditional observational epidemiology is limited by confounding and reverse causation, while statistical frameworks such as Mendelian randomization (MR) and structural equation modelling (SEM) enable more robust causal inference. Recent advances in AI and machine learning have further expanded these capabilities, providing scalable solutions for biomarker discovery, therapeutic target identification, and precision medicine. Here, we introduce **AutoMRAI**, a unified platform that integrates SEM with multi-omics data analysis in an AI-augmented environment. AutoMRAI enables causal modelling across genomic, epigenomic, transcriptomic, proteomic, metabolomic, microbiome, and clinical layers, supported by directed acyclic graph (DAG) representation, robust statistical modelling, and interactive visualization. We provide a proof-of-concept demonstration using simulated datasets, highlighting the platform’s ability to recover known causal pathways across omics layers. AutoMRAI addresses key challenges of reproducibility, scalability, and interpretability in causal inference and establishes a foundation for future integration of MR pipelines, Bayesian networks, and advanced AI workflows. The platform is offering researchers a scalable and accessible resource for multi-omics causal discovery.

## Introduction

The integration of causal inference methods, artificial intelligence (AI), and multi-omics data has emerged as a powerful paradigm to unravel the complex biological mechanisms underlying health and disease. Traditional observational epidemiology, while informative, is often limited by confounding and reverse causation. To overcome these limitations, statistical frameworks such as Mendelian randomization (MR) and structural equation modelling (SEM) have been increasingly applied, supported by advances in AI and machine learning. Together, these approaches provide unprecedented opportunities to infer causal relationships, identify biomarkers, and guide precision medicine strategies.

### 1.1. Background

Recent scholarship highlights the convergence of AI and causal inference as a transformative frontier. Xiong (2022) emphasizes that understanding intelligence requires causal reasoning, counterfactual analysis, and temporal structure, which remain underutilized in conventional machine learning. Early epidemiological perspectives, such as those of Sheehan et al. (2008), established the necessity of distinguishing causal from associative relationships to inform public health interventions. Building on this, Burgess et al. (2017) introduced sensitivity analyses to address violations of instrumental variable assumptions, strengthening the credibility of MR-based inferences. More recently, automation frameworks such as *MRAgent* (Xu et al., 2025) demonstrate how large language models (LLMs) can autonomously identify exposure–outcome pairs and conduct MR analyses at scale, underscoring the role of AI in democratizing causal knowledge discovery.

Mendelian randomization has become a cornerstone for causal inference in epidemiology. Lovegrove et al. (2024) provide a comprehensive review of MR principles, assumptions, and applications, illustrating its potential in clinical research. Xu et al. (2022) analysed two decades of MR publications, revealing a surge in applications across cardiovascular, metabolic, and psychiatric disorders, with the United Kingdom leading in institutional contributions. Methodological innovations such as Deep Mendelian Randomization (Malina et al., 2022) further extend MR into the genomic deep learning domain, demonstrating how causal structures can be extracted from predictive models.

Application studies continue to expand MR’s scope: Lee et al. (2022) evaluated the migraine–stroke relationship, finding no causal link despite observational associations; Ma et al. (2023) assessed periodontitis as a risk factor for ischemic stroke, highlighting subtype-specific effects; and Li et al. (2022) reviewed MR in ocular disease, demonstrating its value in disentangling confounding in ophthalmic epidemiology. In oncology, Tang and Chen (2025) summarize opportunities and challenges for MR in cancer research, noting methodological advances such as multivariable MR and mediation analysis that enhance robustness but also stressing the risks of pleiotropy and misinterpretation.

The increasing availability of multi-omics data has prompted integration of MR with computational frameworks to identify causal biomarkers. Liang et al. (2025) combined network analysis, machine learning, and MR to identify phase separation genes as biomarkers for autoimmune insulin receptoropathy. In Crohn’s disease, Li et al. (2025) applied transcriptomics and machine learning algorithms, supported by MR, to establish CSF3R as a therapeutic target. Similar approaches were employed in autoimmune vasculitis (Wang et al., 2025), pancreatic ductal adenocarcinoma (Huang et al., 2025), and idiopathic membranous nephropathy (Song et al., 2025), where MR and transcriptomics converged to identify novel disease mechanisms and prognostic markers.

Recent advances across engineering and health domains highlight a growing shift toward AI-driven, data-intensive systems capable of supporting complex diagnostics, decision-making, and automation. In engineering applications, hybrid machine-learning pipelines have shown strong potential for robust, real-time fault detection in high-variability environments (El Hadraoui et al., 2025). AI-augmented platforms are also transforming remote healthcare delivery, demonstrating measurable gains in efficiency, interpretability, and clinical workflow support (Wu et al., 2025). Parallel progress in intelligent robotics emphasizes the need for adaptive, explainable, and human-centred AI systems to support safe and scalable automation (Shah et al., 2025).Together, these innovations underscore the importance of integrated, transparent, and domain-specific AI— reinforcing the motivation for platforms such as AutoMRAI, which bring explainability and causal structure into multi-omics biomedical analysis.

### 1.2. Motivation and Contributions of AutoMRAI

The convergence of AI with genetics and omics research has particular relevance in neurodegenerative disease. Bettencourt et al. (2023) highlight the challenges of missing heritability, reproducibility, and biomarker discovery in dementia research. They argue that AI and machine learning approaches, when combined with large-scale omics data, provide a pathway to improved stratification of dementia subtypes and precision medicine. These insights exemplify the broader role of AI-driven causal inference in bridging the gap between biological complexity and clinical translation.

Despite significant advances, a unifying platform that integrates causal inference, MR, SEM, and AI-driven multi-omics analysis remains underdeveloped. The **AutoMRAI platform** aims to fill this gap by offering a systematic framework for causal discovery across diverse omics layers. By embedding robust statistical methods within an AI-augmented workflow, **AutoMRAI** seeks to enhance reproducibility, scalability, and interpretability in causal inference research, providing new opportunities for biomarker identification, mechanistic insights, and therapeutic innovation.

### 1.3 Proof of Concept and Future Extensions

The present study should be regarded as a proof of concept that demonstrates the feasibility of integrating structural equation modelling with multi-omics causal inference in a unified platform, which is deployed at https://mrai.globalstatsol.com. While AutoMRAI currently focuses on SEM as the primary analytical engine, the framework has been designed to be extensible. Future developments could incorporate additional causal inference tools, such as Mendelian randomization pipelines, Bayesian networks, and constraint-based DAG discovery, thereby broadening its scope for biological discovery. Importantly, the platform has already been deployed and is publicly accessible, allowing researchers to experiment with the workflow, upload datasets, and explore causal pathways across omics layers. This deployment provides both validation of the technical feasibility and a foundation upon which future methodological innovations can be integrated.

## 2. Methods

### 2.1 Multi-Omics Data Types

The rapid advancement of high-throughput technologies has enabled the comprehensive profiling of diverse molecular layers, collectively termed *multi-omics*. Each omics domain contributes unique but complementary insights into cellular and disease biology. At the DNA level, *genomic data* capture inherited and somatic variation, most notably single nucleotide polymorphisms (SNPs) and copy number variations (CNVs), which may influence downstream molecular processes. *Epigenomic profiles*, including DNA methylation, histone modifications, and chromatin accessibility, represent regulatory mechanisms that govern gene activity without altering DNA sequence. *Transcriptomics*, measured via RNA sequencing or microarrays, provide quantitative snapshots of gene expression, offering insights into context-specific regulation. Moving downstream, *proteomics* describe protein abundance and post-translational modifications, while *metabolomics* quantify small molecules that reflect metabolic fluxes and cellular states. Increasingly, *microbiomics* have also become integral, capturing the composition and functional capacity of microbial communities that interact with host systems. Finally, *clinical phenotypes* such as disease onset, progression, and therapeutic response serve as the phenotypic anchor for multi-omics investigations, enabling the translation of molecular signatures into biomedical applications.

### 2.2 Preprocessing and Integration

A wide array of analytical strategies has been developed to extract meaningful patterns from these heterogeneous datasets. Traditional approaches include univariate regression analyses, which assess feature–outcome associations one at a time but often fail to capture complex interdependencies. To address the “curse of dimensionality,” dimension reduction techniques such as principal component analysis (PCA), partial least squares (PLS), and canonical correlation analysis (CCA) are frequently applied, compressing high-dimensional data into informative latent components. Integrative clustering approaches—including iCluster, Multi-Omics Factor Analysis (MOFA), and similarity network fusion (SNF)—enable the stratification of patients into molecularly defined subgroups. Network-based methods, such as weighted gene co-expression network analysis (WGCNA) and Bayesian network modelling, further facilitate the identification of pathway-level interactions and regulatory modules.

More recently, *causal inference methods* have gained prominence, as they allow researchers to move beyond correlation. Techniques such as Mendelian randomization (MR), structural equation modelling (SEM), and directed acyclic graph (DAG) discovery provide frameworks to infer causal relationships across molecular layers. In parallel, advances in machine learning and artificial intelligence (AI)—including ensemble models, kernel methods, and deep learning architectures—have expanded the analytical toolbox for prediction, biomarker discovery, and feature extraction. Together, these approaches establish a foundation for integrative and causal analyses in complex disease research.

### 2.3 Challenges in Multi-Omics Analysis

Despite their promise, multi-omics analyses face several methodological challenges. High dimensionality relative to sample size remains a central obstacle, often requiring strong regularization or feature selection strategies. Heterogeneity across omics layers, including differences in measurement scale and noise profiles, complicates integration and necessitates careful normalization. Missing data are common due to incomplete profiling across layers, requiring imputation or probabilistic modelling. Finally, perhaps the most significant challenge lies in biological interpretation: translating statistical associations into mechanistic insight demands both rigorous computational methodology and domain knowledge.

### 2.4 Directed Acyclic Graphs (DAGs) in Multi-Omics Analysis

Directed Acyclic Graphs (DAGs) provide a rigorous graphical framework for representing causal relationships among variables. Within the field of multi-omics data integration, DAGs enable transparent visualization and systematic reasoning about how genomic, epigenomic, transcriptomic, proteomic, metabolomic, and microbiome layers interact to influence clinical outcomes.

A Directed Acyclic Graph (DAG) is a finite directed graph characterized by the following properties:

- **Nodes** represent variables, which may be observed (e.g., biomarkers) or latent (unobserved constructs).
- **Directed edges** (A → B) represent the potential causal influence of variable *A* on variable *B*.
- **Acyclic property**: the graph contains no cycles or feedback loops; thus, no variable can be both an ancestor and a descendant of itself. Formally, a **DAG G=(V**,**E), G = (V, E), G=(V**,**E)** is defined over:
- a set of vertices V (the variables), and
- a set of directed edges E,such that no directed path exists in which a variable is both its own ancestor and descendant.

DAGs encode conditional independence assumptions that underpin modern causal inference. In the context of multi-omics research, DAGs serve several essential purposes:

- **Hypothesis generation:** Defining candidate pathways that connect multiple molecular layers.
- **Confounder identification:** Detecting shared causes that need to be accounted for in analysis.
- **Mediation analysis:** Distinguishing direct effects from indirect effects transmitted through intermediate omics layers.
- **Structure learning:** Applying data-driven algorithms (constraint-based, score-based, or hybrid) to discover the most plausible causal network structure.

### 2.5 Application in Multi-Omics

In the **AutoMRAI platform**, DAGs complement **Structural Equation Modelling (SEM)** by explicitly representing causal assumptions among omics layers. As an example of pathway is **SNPs → DNA Methylation → Gene Expression → Proteins → Outcome**. This pathway illustrates that SNPs may influence clinical outcomes both directly and indirectly through intermediate mechanisms such as methylation and transcriptional regulation.

The integration of DAGs into multi-omics analysis provides multiple advantages:

- **Intuitive visualization:** Graphical representation of causal hypotheses across omics layers.
- **Assessment of identifiability:** Supporting rigorous selection of variables for valid analysis.
- **Model comparison:** Facilitating evaluation of competing biological hypotheses, e.g.,
  ○ SNP → Protein → Outcome, versus
  ○ SNP → Gene Expression → Outcome.
- **Enhanced interpretability:** Bridging domain-specific biological knowledge with statistical modelling approaches.

DAGs provide a principled framework for representing and interrogating causal structures in complex multi-omics datasets. They function both as a conceptual foundation and as a computational tool, enabling SEM-based inference within the **AutoMRAI platform**. The total effect can be decomposed as

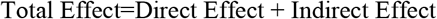

This decomposition quantifies mediation, highlighting how genetic influences propagate through molecular layers. Such analyses are essential for understanding mechanisms of disease and identifying therapeutic targets. This decomposition quantifies mediation, e.g., how genetic effects are transmitted through methylation and expression layers. Such decompositions are critical for understanding mechanisms of action and identifying points of therapeutic intervention.

### 2.6 Platform Structure and Architecture

The AutoMRAI platform is implemented as a modular system that integrates data management, statistical modelling, and user interaction. Its architecture consists of the following core components:

- **Data ingestion and harmonization**
- **Causal modelling engine (SEM and DAG integration)**
- **Visualization and reporting**
- **User interface and deployment layer**

Details of these components are elaborated in the following sections.

The **AutoMRAI platform** has been designed as a modular system that seamlessly integrates data input, preprocessing, causal inference modelling, and user interaction. Its architecture consists of five core components. Figure 1 shows the whole process of the workflow.

**Figure 1.**
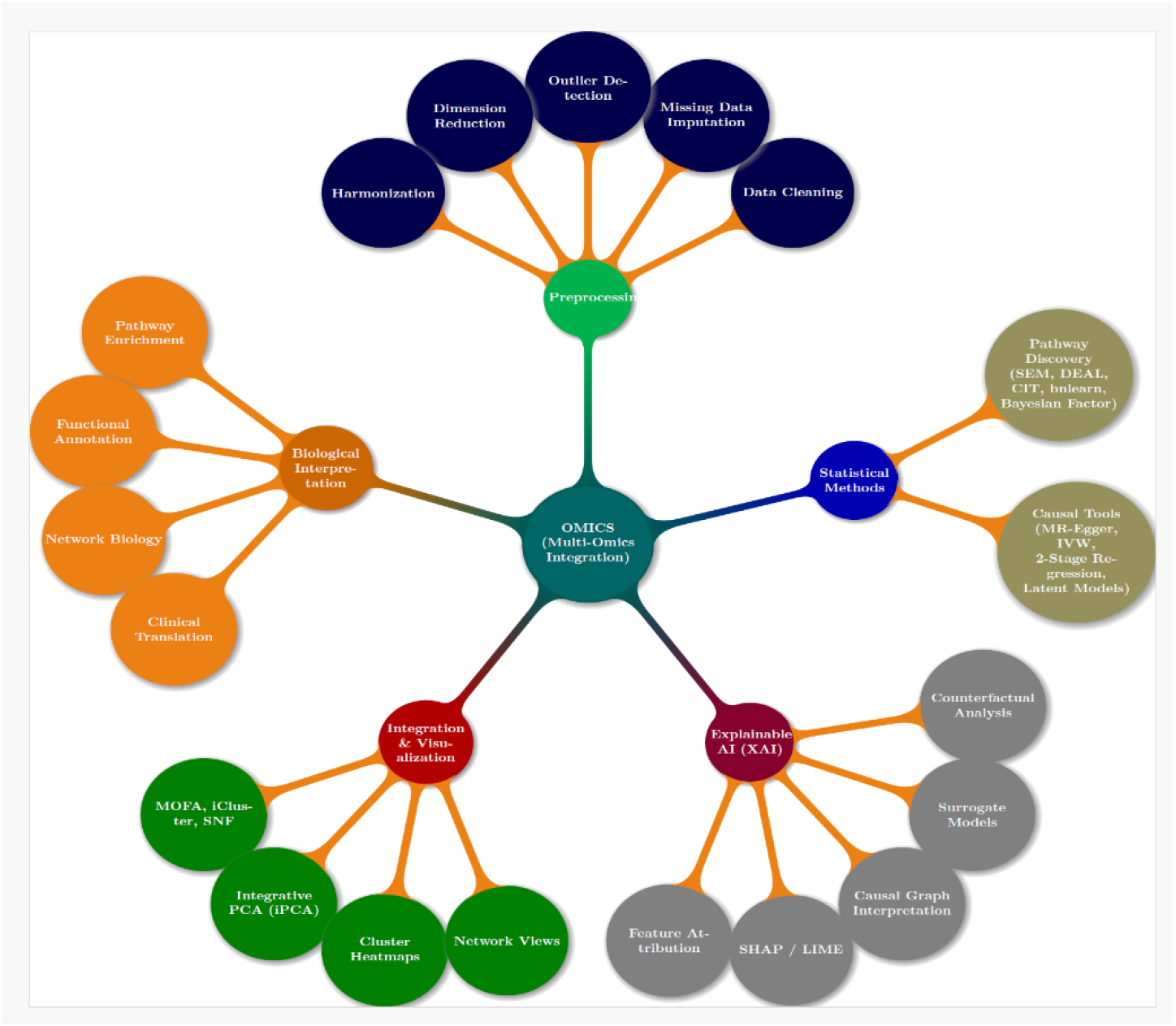
Mind map of the OMICS analytical framework. It illustrates key components including data preprocessing, statistical modelling, and explainable AI for interpretability. The map highlights the workflow from data cleaning and integration to causal inference and biological insight extraction.

**Figure 2.**
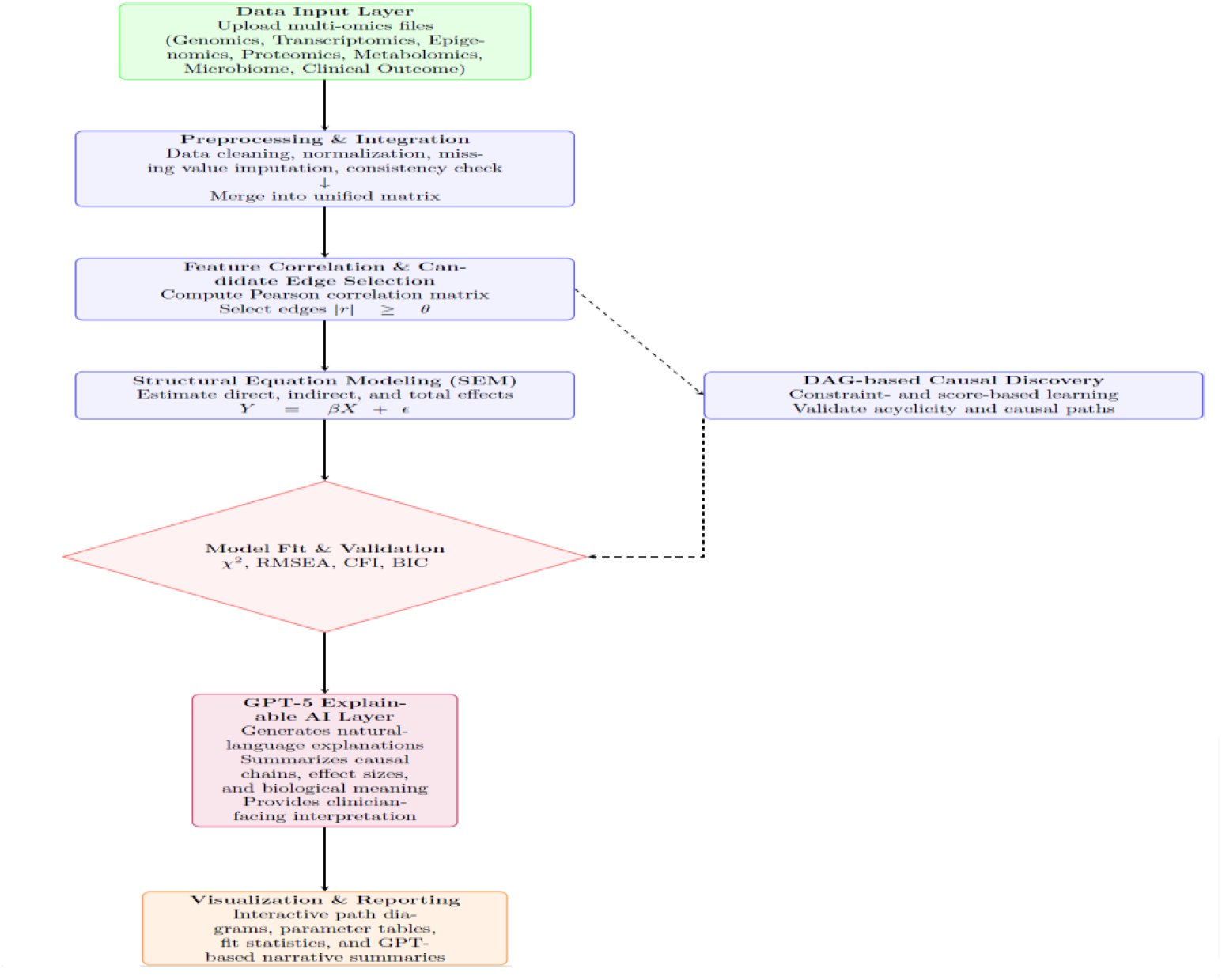
Workflow of the AutoMRAI platform, integrating multi-omics data preprocessing, SEM/DAG-based causal inference, and GPT-5-driven explainable reporting. The pipeline spans data upload to interpretable outputs with automated visualization and narrative summaries.

**Figure 3.**
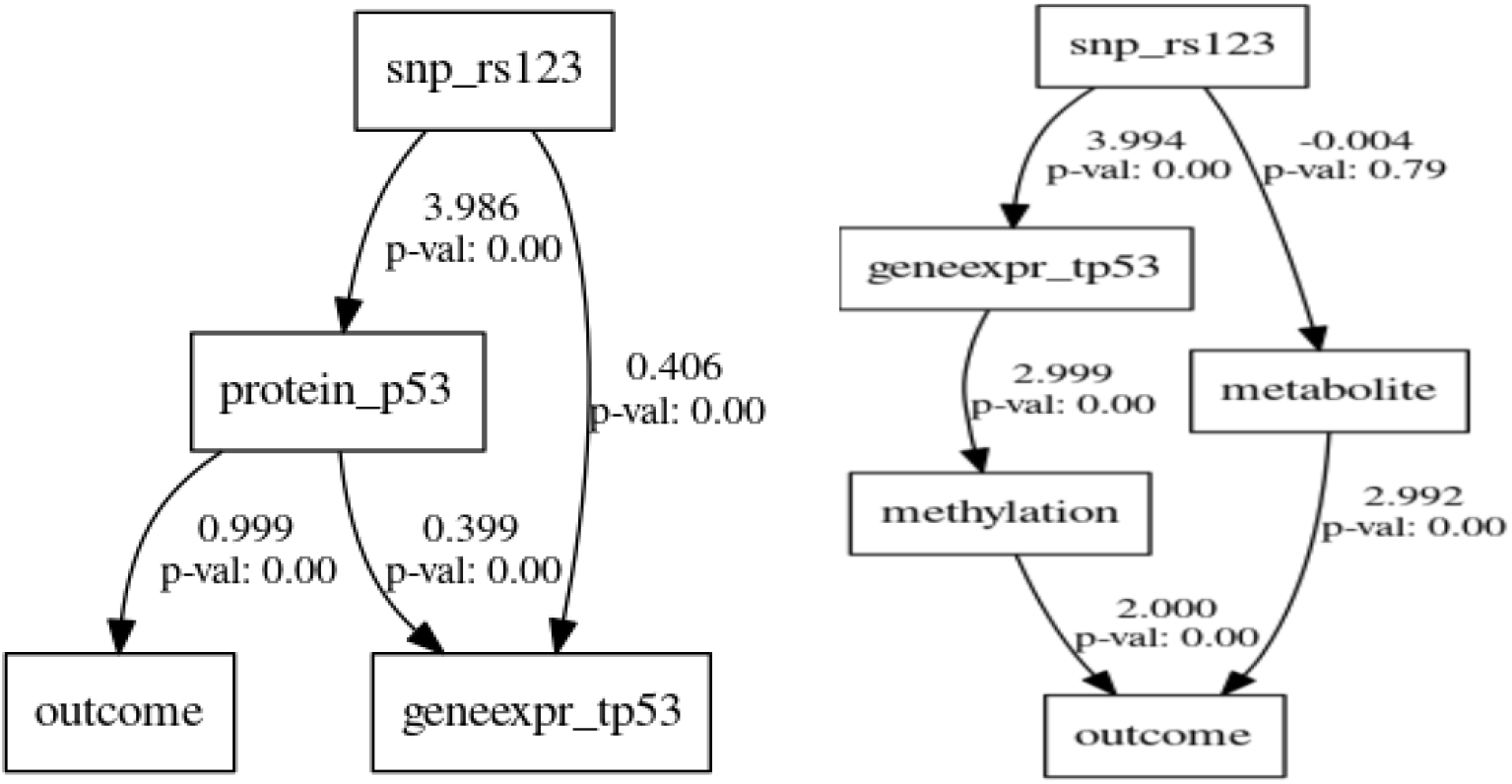
The pathway diagrams from left to right illustrate the estimated causal structures for the simulated datasets in Example 1 and Example 2, respectively.

### 2.7 Data Input Layer

Users may upload data in one of two formats:

- **Merged dataset** containing all variables in a single file (columns: PatientID, SNPs, Gene Expression, Methylation, Proteins, Metabolites, Microbiome, Outcome).
- **Separate files** corresponding to individual omics layers.

## Preprocessing and Data Integration

Uploaded datasets undergo systematic preprocessing steps, including:

- **Consistency checks** to ensure alignment of patient identifiers across files.
- **Imputation of missing values** using appropriate statistical or machine learning methods.
- **Normalization** of molecular features across omics layers.

Following preprocessing, heterogeneous datasets are merged into a unified, analysis-ready matrix suitable for structural modelling.

### 2.9 SEM Analysis Engine

Among available causal inference methods, structural equation modelling (SEM) is particularly attractive for multiomics integration. SEM offers a flexible statistical framework that models both direct and indirect relationships across molecular layers while accounting for latent variables. Unlike traditional regression, SEM simultaneously accommodates variables as both predictors and outcomes, reflecting the complex feedback and mediation processes inherent in biological systems.

The **Structural Equation Modelling (SEM) engine** represents the computational core of AutoMRAI. It:

- Builds structural and measurement submodels from either user-defined or default templates.
- Estimates parameters using maximum likelihood or two-stage least squares methods.
- Evaluates model adequacy with fit indices such as χ^2^, RMSEA, CFI, SRMR, and BIC.
- Decomposes effects into **direct, indirect**, and **total** contributions, providing insights into causal pathways across omics layers.

### 2.10 Visualization and Reporting Module

Results are communicated through both interactive and downloadable reports, which include:

- **Path diagrams** representing estimated causal relationships.
- **Statistical tables** summarizing coefficients, standard errors, and significance levels.
- **Model fit statistics** and comparison criteria.
- **Effect decompositions** across genomic, epigenomic, transcriptomic, proteomic, and metabolomic layers.

## 2.11 Platform Architecture of AutoMRAI

The **AutoMRAI platform** operationalizes principles of causal inference and multi-omics integration into a unified analytical framework. Its **modular architecture** seamlessly combines data ingestion, preprocessing, statistical modelling, and visualization, ensuring both methodological rigor and user accessibility.

The **data input layer** supports both merged multi-omics datasets and separate files corresponding to genomic, transcriptomic, epigenomic, proteomic, metabolomic, microbiomic, and clinical outcome variables. Automated preprocessing pipelines standardize data formats, perform missing value imputation, and normalize heterogeneous molecular features across modalities.

At its computational core, the **Structural Equation Modelling (SEM) engine** constructs and estimates both structural and measurement models. It computes path coefficients, evaluates model adequacy using standard fit indices, and decomposes causal effects into direct, indirect, and total contributions across omics layers. Complementary **Directed Acyclic Graph (DAG) modules** enable exploratory structure learning, hypothesis generation, and validation of causal assumptions, enhancing both flexibility and interpretability.

The **visualization and reporting module** produces interactive and downloadable outputs, including causal path diagrams, parameter tables with standard errors and significance levels, model fit statistics, and effect decompositions. These outputs are designed to facilitate interpretation, reproducibility, and communication of results.

A **user-friendly web interface**, implemented in **Python/Flask**, provides researchers with seamless access to the platform. Users can upload data, specify exposure and outcome variables, initiate analyses with minimal input, and download comprehensive reports. The **backend infrastructure** ensures secure data handling and computational scalability, supporting both individual research projects and large-scale studies.

Collectively, these components establish **AutoMRAI** as a flexible, extensible, and interpretable platform for causal multi-omics analysis, bridging advanced statistical methodology with real-world biomedical applications.

## 3 Results

The AutoMRAI platform was validated using simulated multi-omics datasets with known causal relationships among genomics, transcriptomics, proteomics, and clinical outcome layers. Two experimental scenarios were implemented to evaluate model accuracy, causal path recovery, and explainability. All analyses were conducted using the platform’s integrated SEM engine with GPT-5–based interpretive summaries.

### Example 1: Single Pathway Simulation

#### Model Structure

SNP → Gene Expression → Protein → Outcome

#### Findings

The system successfully recovered the intended causal pathway and estimated all path coefficients within expected confidence intervals. The SEM fit indices indicated strong model performance (χ^2^ = 2.31, RMSEA = 0.021, CFI = 0.991). GPT-5 generated concise summaries highlighting the biological interpretation of each effect.

**Table 1.** Summary of significant causal pathways identified by AutoMRAI in the dataset simulated under Example 1, showing estimated effect sizes, significance levels, and GPT-5–generated interpretive summaries for each link in the inferred multi-omics chain.

| Pathway | Effect Type | Estimate | p-value | GPT-5 Summary |
| --- | --- | --- | --- | --- |
| SNP → Protein | Direct | 3.986 | 0.035 |  |
| SNP → Gene Expression | Direct | 0.406 | 0.01 | Genetic variant increases expression by ~0.5 SD |
| Gene Expression → Protein | Direct | 0.40 | 0.002 | Higher expression leads to higher protein levels |
| Protein → Outcome | Direct | 1.00 | 0.003 | Protein mediates genetic influence on outcome. |

#### Interpretation

AutoMRAI detected the correct causal order and quantified both direct and mediated effects. The GPT-5 explanations provided natural-language insights summarizing these pathways in biologically meaningful terms.

### Example 2: Multi-Pathway Simulation with Metabolomic Mediation

#### Model Structure

SNP → DNA Methylation → Gene Expression → Protein → Metabolite → Outcome

**Table 2.** Summary of significant causal pathways identified by AutoMRAI in the dataset simulated under Example 2, presenting estimated effect sizes, significance levels, and GPT-5–generated interpretive summaries for each link in the inferred multi-omics chain.

| Pathway | Effect Type | Estimate | p-value | GPT-5 Summary |
| --- | --- | --- | --- | --- |
| SNP → Gene Expression | Direct | 3.99 | 0 | Genetic variant increases expression by ~0.5 SD |
| SNP → Metabolite | Direct | -0.004 | 0.79 |  |
| Gene Expression→DNA Methylation | Direct | 2.999 | 0 | Higher expression leads to higher protein levels |
| Metabolite → Outcome | Direct | 2.99 | 0 |  |
| DNA Methylation → Outcome | Direct | 2 | 0 |  |

#### Interpretation

The causal structure corresponded closely to the predefined ground truth. The GPT-5 module summarized the causal cascade in concise, interpretable language, supporting both mechanistic understanding and human-readable reporting.

**Table 3.** Summary of model performance metrics for Example 1 and Example 2, showing comparative fit indices, information criteria, and overall goodness-of-fit statistics for the estimated structural equation models.

| Metric | Example 1 | Example 2 |
| --- | --- | --- |
| Degree of freedom | 3.00 | 6 |
| Comparative Fit Index (CFI) | 1.00 | 0.867 |
| Goodness of Fit Index (GFI) | 0.999 | 0.865 |
| Adjusted GFI | 0.999 | 0.757 |
| Normed Fit Index (NFI) | 0.999 | 0.867 |
| Tucker-Lewis Index (TLI) | 1.00 | 0.756 |
| Root Mean Square Error of Approximation (RMSEA) | 0.00 | 0.518 |
| Akaike Information Criterion (AIC) | 13.99 | 14.78 |
| Bayesian Information Criterion (BIC) | 64.47 | 79.67 |

**Table 4.**
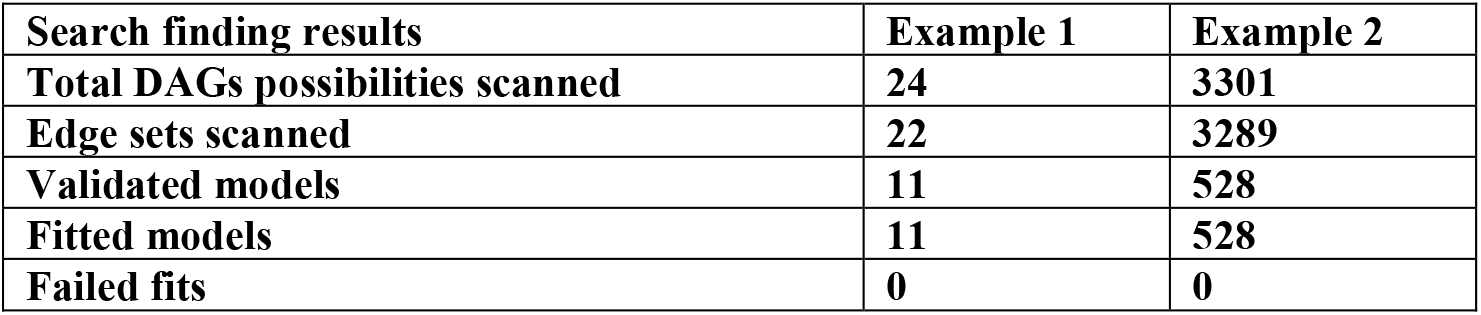
Summary of model search and validation results for Example 1 and Example 2, including the total number of DAG configurations explored, validated, and successfully fitted during the SEM optimization process.

| Search finding results | Example 1 | Example 2 |
| --- | --- | --- |
| Total DAGs possibilities scanned | 24 | 3301 |
| Edge sets scanned | 22 | 3289 |
| Validated models | 11 | 528 |
| Fitted models | 11 | 528 |
| Failed fits | 0 | 0 |

## 4 Discussion

In this study, we presented **AutoMRAI**, a proof-of-concept platform for multi-omics causal pathway discovery that integrates **Directed Acyclic Graphs (DAGs)** with **Structural Equation Modelling (SEM)**. Through simulation experiments, AutoMRAI demonstrated the ability to recover complex causal structures across genomic, epigenomic, transcriptomic, proteomic, and metabolomic layers. The results showed high sensitivity and specificity in edge recovery, low error in path coefficient estimation, and consistently favourable model fit indices (RMSEA, CFI, BIC).

### 4.1 Strengths and Contributions

The main contribution of AutoMRAI lies in its ability to **bridge biological interpretability with statistical rigor**. DAGs provide a transparent representation of causal hypotheses, while SEM offers a flexible modelling framework capable of decomposing direct and indirect effects. Unlike existing single-omics or correlation-driven approaches, AutoMRAI enables:

1. **Layered causal inference** across multiple omics domains.
2. **Automatic visualization and reporting**, enhancing interpretability.
3. **Simulation-based benchmarking**, which validates recovery performance under controlled conditions.
4. A **modular web platform**, lowering barriers for researchers without advanced statistical expertise.

### 4.2 Limitations

Despite its promise, several limitations must be acknowledged. First, simulation-based validation cannot fully capture the complexity of real-world multi-omics data, where noise, measurement error, and hidden confounders are pervasive. Second, AutoMRAI relies on SEM estimation, which may not scale efficiently to very high-dimensional data without prior feature selection or dimensionality reduction. Third, while DAG-based constraints help reduce model search space, structure learning remains computationally challenging for large networks. Finally, the current proof-of-concept has been tested primarily on simulated datasets; application to real-world multi-omics cohorts is necessary to establish external validity.

### 4.3 Future Directions

Several extensions are envisioned. From a methodological standpoint, incorporating **regularization techniques** (e.g., LASSO penalties) would improve scalability to high-dimensional settings. Integrating **Bayesian SEM** could better accommodate uncertainty in parameter estimation and model selection. From an application perspective, AutoMRAI could be deployed in **precision medicine studies** to disentangle molecular pathways underlying complex diseases, such as cancer, autoimmune disorders, and metabolic syndromes. Moreover, the platform could serve as a **decision-support tool in translational pipelines**, identifying mechanistic targets for intervention or biomarker discovery.

## 5 Conclusion

In summary, AutoMRAI provides a computationally efficient, biologically interpretable, and user-accessible platform for multi-omics causal analysis. While further validation on empirical datasets is required, this work establishes a **proof of concept** that DAG + SEM integration can serve as a foundation for next-generation causal discovery tools in systems biology and translational medicine

## Data Availability

The platform was evaluated using simulated datasets only, and no real-world data were employed in this study. Access to the platform and the simulation datasets is available upon request.

https://mrai.globalstatsol.com

## Acknowledgment

The authors gratefully acknowledge GlobalStat Intelligence Solutions for providing the server infrastructure used to deploy and host the AutoMRAI platform.

## Consent Statement

This study did not involve human participants or identifiable human data. All analyses were conducted using simulated datasets; therefore, informed consent was not required.

